# Prevalence and predisposing factors of Hookworm infection, and Anemia among Pregnant women attending antenatal care in Dawuro zone, southwest Ethiopia, Institution-based cross-sectional study

**DOI:** 10.1101/2025.06.28.25330414

**Authors:** Getachew Teshome, Berhanu Effa, Asrat Chernet, Temesgen Woldemedhin, Mengistu Geribo

## Abstract

Hookworm infection and anemia are significant public health concerns in developing countries, particularly among pregnant women in low-resource settings. These conditions can lead to adverse maternal and fetal outcomes.

**Objectives:** This study aimed to assess the prevalence and identify predisposing factors of Hookworm infection and anemia among pregnant women attending antenatal care in Dawuro Zone, Southwest Ethiopia.

**Methods:** Institution-based cross-sectional study was conducted from December 2022 to March 2023, among pregnant women attending antenatal clinics in selected public health institutions. A total of 367 participants were selected using systematic random sampling. Data were collected through interviewer-administered questionnaires. Stool samples were collected and examined by direct wet mount and Formol-ether concentration methods to detect Hookworm, and hemoglobin levels were measured using a HemoSmart analyzer. Logistic regression analysis was used to identify factors associated with Hookworm infection and anemia.

**Result:** The total of 367 pregnant women of different age groups and different trimester of pregnancy were diagnosed for presence of Hookworm infection and anemia. The response rate was 100%. The prevalence of Hookworm and anemia among pregnant women was found to be 10.6% and 31.9% respectively with mean hemoglobin concentration of 11.334±1.92 gm/dl. Hookworm infection and anemia were significantly associated with walking barefoot (AOR=2.89, 95% CI: 1.27–6.60), poor sanitation practices (AOR=2.89, 95% CI: 1.24–6.74), inadequate supplementation taken (AOR=4.59, 95% CI: 1.8–11.5), and low household income (AOR=7.0, 95% CI: 3.27–14.98).

## 1. INTRODUCTION

Hookworm infection, caused by the nematodes Necator americanus and Ancylostoma duodenale, remains a major public health challenge in low- and middle-income countries. Transmission typically occurs through skin penetration by infective larvae from contaminated soil, especially among individuals who walk barefoot [1]. The infection is widespread in tropical regions due to favorable climates and poor sanitation [2]. Though often asymptomatic, Hookworm can lead to anemia, malnutrition, and other complications, particularly in vulnerable groups such as pregnant women [3]. Anemia, defined by the World Health Organization (WHO) as a hemoglobin (HGB) level below 11 g/dL during pregnancy, impairs oxygen transport and increases the risk of maternal and fetal complications, including preterm birth, low birth weight, and maternal mortality [4-5]. The burden of anemia is most severe in sub-Saharan Africa, where both nutritional deficiencies and parasitic infections contribute significantly [6]. Globally, an estimated 44 million pregnant women are infected with Hookworm, contributing to significant economic losses and health impacts [7]. Based on recommended HGB concentrations thresholds (<10.5 g/dl in second trimester of pregnancy, <11 g/dl in first and third trimester of pregnancy, and under-5 children, <11.5 g/dl in pre-school children, <12 g/dl in adolescents, and non-pregnant women and <13 g/dl in men), the WHO estimates that 2 billion people are anemic [8-9]. Despite deworming being included in Ethiopia’s antenatal care (ANC) services, its utilization remains low, especially among rural, uneducated, and low-income women. In the Dawuro Zone of Southwest Ethiopia, data on Hookworm infection and anemia among pregnant women is limited. This study aims to assess their prevalence and identify key predisposing factors to enlighten public health interventions.

## 2. METHODS AND MATERIALS

### 2.1. Study area, period and Study Design

An institution-based cross-sectional study was conducted from December 2022 to March 2023 in the Dawuro Zone, located approximately 515 km southwest of Addis Ababa, Ethiopia. The zone is bordered by Gamo Gofa to the south, Konta to the west, the Gojeb River and Oromia region to the north, Hadiya and Kembata Tembaro to the northeast, and Wolaita zone and the Omo River to the east and southeast. Dawuro Zone comprises 10 woredas and two administrative towns, with a predominantly rural population. According to the 2007 Ethiopian census, the total population was 684, 702. The area features diverse topography including mountains, plateaus, lowlands, and gorges. The local economy is primarily based on agriculture, with major crops including maize, enset, sorghum, and root vegetables, alongside cattle rearing. Health infrastructure includes one general hospital, two district hospitals, 21 health centers, 178 health posts, and multiple private clinics and drug stores, serving both rural and urban kebeles.

### 2.2. Study Population, Sample size Determination and Sampling Technique

The source population consisted of all pregnant women attending ANC clinics in the Dawuro Zone. The study population included those attending selected health facilities that consented to participate. Pregnant women with recent blood loss and severe dehydration were excluded. The sample size was calculated using a single population proportion formula, assuming a 95% confidence level, 5% margin of error, and a 32% prevalence of Hookworm infection based on a prior study in Dembecha District, Northwest Ethiopia. After adding a 10% non-response rate, the final sample size was 367. Health facilities were selected through random sampling, including one general hospital, one district hospital, and three health centers. The sample was proportionally allocated based on the ANC client load at each facility. Participants were selected using systematic random sampling, with the sampling interval (K) calculated by dividing the total number of ANC attendees by the sample size. The first participant was chosen randomly by lottery, followed by every K^th^ attendee.

### 2.3. Study Variables

The outcome variables were the prevalence of Hookworm infection and anemia. Independent variables included socio-demographic characteristics (age, marital status, education, residence, occupation, household income, etc.), obstetric history (gestational age, gravidity, parity, history of abortion, first pregnancy, etc.), and other potential risk factors

### 2.4. Laboratory Works

#### 2.4.1. Stool Sample Collection and Examination

After obtaining informed consent, socio-demographic and clinical data were collected via a structured questionnaire administered through face-to-face interviews in the local language by trained nurses and midwives; each participant was given a unique code, which was used on both the questionnaire and lab request form. Participants were asked to provide approximately 20 g of stool in a clean, dry, leak-proof container. Samples were examined using direct wet mount and Formol-ether Concentration methods.

##### Direct Wet Mount Microscopy

A fresh stool sample (about 2mg of stool) was placed on two slides with a wooden applicator stick, emulsified with a drop of physiological saline (0.85%), covered with cover slides, and examined under a microscope using a 10x objective of microscope for the presence of Hookworm ova.

##### Formol-Ether Concentration

This test was performed by mixing thoroughly around 1 g of feces in 3-4 ml of 10% formaldehyde in a glass container. Two layers of gauze were placed in a funnel, and the contents were strained into a 15 ml centrifuge tube. Then, additional 3 ml of 10% formaldehyde and 3 ml of ether were added. The solution was mixed well and centrifuged at 1000 revolution for 3 min. The supernatant was removed, and two slides were prepared from the sediment and finally examined with a 10x objective of the microscope. For those participants who were positive for any intestinal parasites in direct wet mount or concentration technique were treated by recommended drugs based on WHO guidelines.

#### 2.4.2. Blood Sample Collection and Hematological Analysis

Capillary blood was collected aseptically from the fingertip and analyzed using a HemoSmart hemoglobinometer, following manufacturer instructions. After calibrating the HemoSmart hemoglobinometer, the strip was inserted, one drop (1μl) of blood was applied, and the HGB levels was reported in g/dl and hematocrit (HCT) in percent within 5 minutes.

Thick and thin blood smears were also prepared on clean slide, stained with Giemsa, and examined for hemoparasites and red blood cell morphology to assess anemia type. Anemic participants were treated per WHO recommendations. The human immunodeficiency virus (HIV) status of the respondents was taken from clients’ records and history of chronic illness was also assessed by asking respondents and those on medication, advised to use preventive actions, and had follow up visits for such diseases.

### 2.5. Data Analysis and Interpretation

Data were entered into Epidata version 3.1 and exported to SPSS version 22 for analysis. Descriptive statistics were used to summarize participant characteristics. Bivariate logistic regression identified potential associations (P< 0.25), and those variables were entered into multivariate logistic regression to determine independent predictors. Statistical significance was set at P< 0.05.

### 2.6. Data Quality Assurance and Ethical Consideration

Data collectors received training on study objectives and questionnaire content. Daily supervision ensured consistency and completeness. Laboratory procedures followed standard operating procedures (SOPs), and 10% of the samples were randomly re-examined blindly by experienced Medical Laboratory technologists for quality control. Ethical clearance was obtained from the Institutional Review Board (IRB) of Wolaita Sodo University (Ref. No: WSU 41/30/314). Official letters of permission were secured from the Dawuro Zone Health Department and respective Woreda Health Offices.

#### Operational term definition

Prevalence: - is the number of Hookworm and anemia observed during the study period.

Anemia in pregnancy: - Hgb and HCT level are less than cut-off value during each trimester of pregnancy.

Mild Anemia= HGB concentration of 10–10.9 g/dl Moderate Anemia = HGB concentration of 7–9.9 g/dl Severe Anemia = HGB concentration less than 7.0 g/dl

Pregnant woman: A woman whose pregnancy was confirmed by Human chorionic gonadotrophin hormone (HCG) test or abdominal examination and fetoscope at the study health facilities.

## 3. RESULTS

### 3.1. Socio-Demographic Characteristics

A total of 367 pregnant women of different age and different trimesters of pregnancy were participated in the study. The response rate was 100%. Participants ranged in age from 18 to 44 years, with a mean age of 25.25 ± 4.82 years. The majority were aged 20–24 years (38.0 %), followed by 25–29 years (30.0 %). Most were in their second trimester (47.4 %), followed by 43.1% in the third and 9.5% in the first trimester. The average family size was 4.13 ± 1.55, and 60.5% had five or more family members. In terms of residence, 63.5% lived in rural areas. Housewives accounted for the largest occupational group (41.4 %), followed by government employees (25.1 %), farmers (17.2 %), students (8.4 %), and merchants (7.9 %). Most participants were married (99.0 %), belonged to the Dawuro ethnic group (95.4 %), and identified as Protestant Christians (80.0 %). Educationally, 93.5% were literate, and 52.9% reported a monthly family income above 3000 Ethiopian Birr (Table 1).

**Figure 1:**
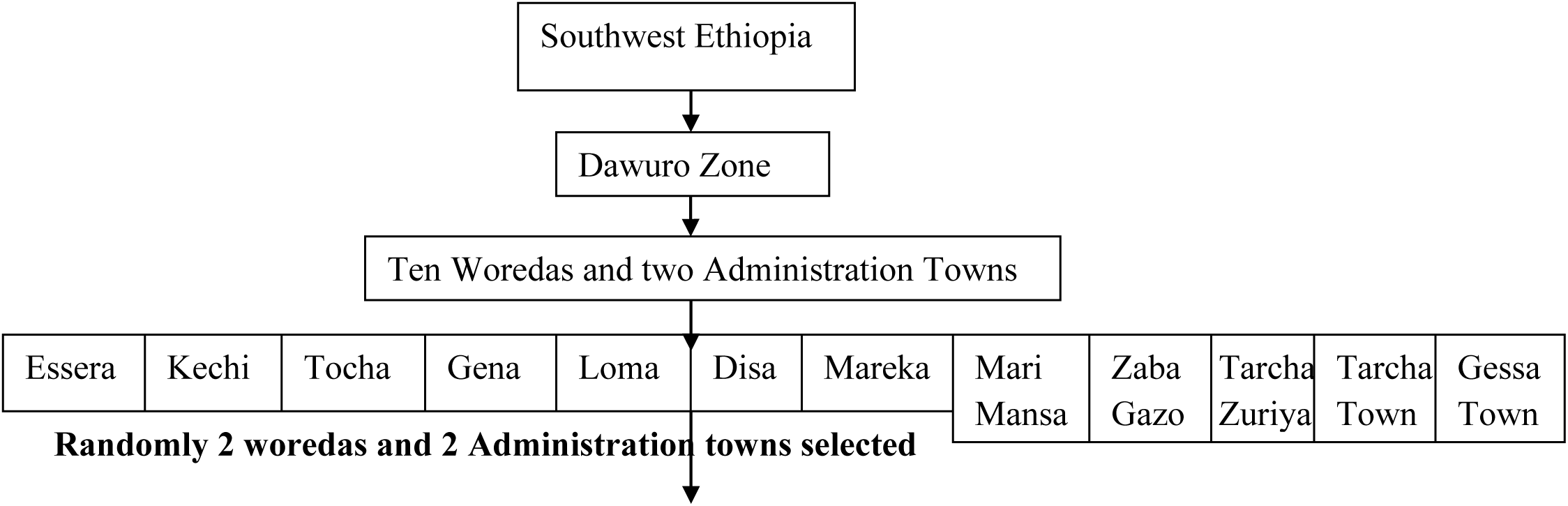
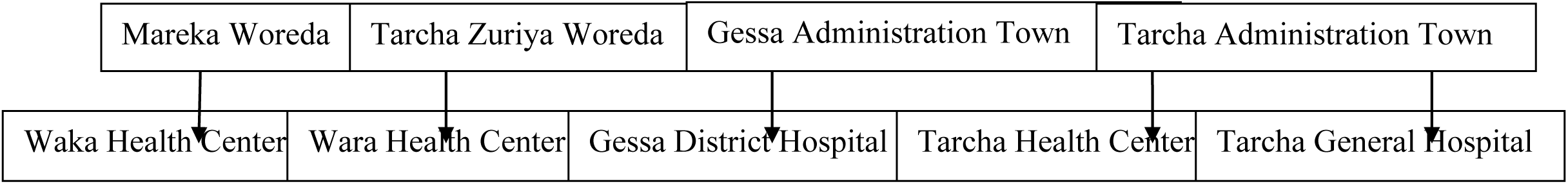
Diagrammatic Representation of the study area, selected woredas and their respective Health facilities of Dawuro Zone, Southwest Ethiopia

**Table 1:** Socio-demographic characteristics among pregnant women attending ANC in Dawuro zone, Southwestern Ethiopia, 2022/23 (n=367).

| Variables | Frequency | Percentage (%) |
| --- | --- | --- |
| **Age in year** |  |  |
| 15-19 | 43 | 12.0 |
| 20-24 | 140 | 38.0 |
| 25- 29 | 112 | 30.0 |
| $\geq 30$ | 72 | 20.0 |
| **Marital Status** |  |  |
| Single | 04 | 1.0 |
| Married | 363 | 99.0 |
| **Ethnicity** |  |  |
| Dawuro | 350 | 95.4 |
| Amhara | 11 | 3.0 |
| Hadiya | 06 | 1.6 |
| **Religion** |  |  |
| Protestant | 294 | 80.0 |
| Orthodox | 68 | 19.0 |
| Catholic | 05 | 1.0 |
| **Family size** |  |  |
| <5 | 145 | 39.5 |
| ≥5 | 222 | 60.5 |
| **Educational status** |  |  |
| No formal education | 24 | 6.5 |
| Literate | 343 | 93.5 |
| **Occupational status** |  |  |
| Housewife | 152 | 41.4 |
| Farmer | 63 | 17.2 |
| Student | 31 | 8.4 |
| Governmental employee | 92 | 25.1 |
| Merchant | 29 | 7.9 |
| **Residence** |  |  |
| Rural | 233 | 63.5 |
| Urban | 134 | 36.5 |
| **Family income** |  |  |
| < 3000 ETB | 173 | 47.1 |
| ≥ 3000 ETB | 194 | 52.9 |

### 3.2. Hygiene and Sanitation-Related Characteristics of the Households

Among the 367 households assessed, 61.3% had their own latrine, while 38.7% lacked access to any latrine. Of those with latrines, 65.1% lacked proper lids, and 57.8% had no hand washing facilities nearby their latrine. A significant proportion of participants (80.7 %) reported not wearing shoes regularly, increasing the risk of Hookworm infection. There were statistically significant associations between both Hookworm infection and anemia with the following hygiene-related factors: lack of a private latrine, open defecation, absence of a latrine lid, and lack of hand washing facilities and barefoot walking (p < 0.05) (Table 2).

**Table 2:**
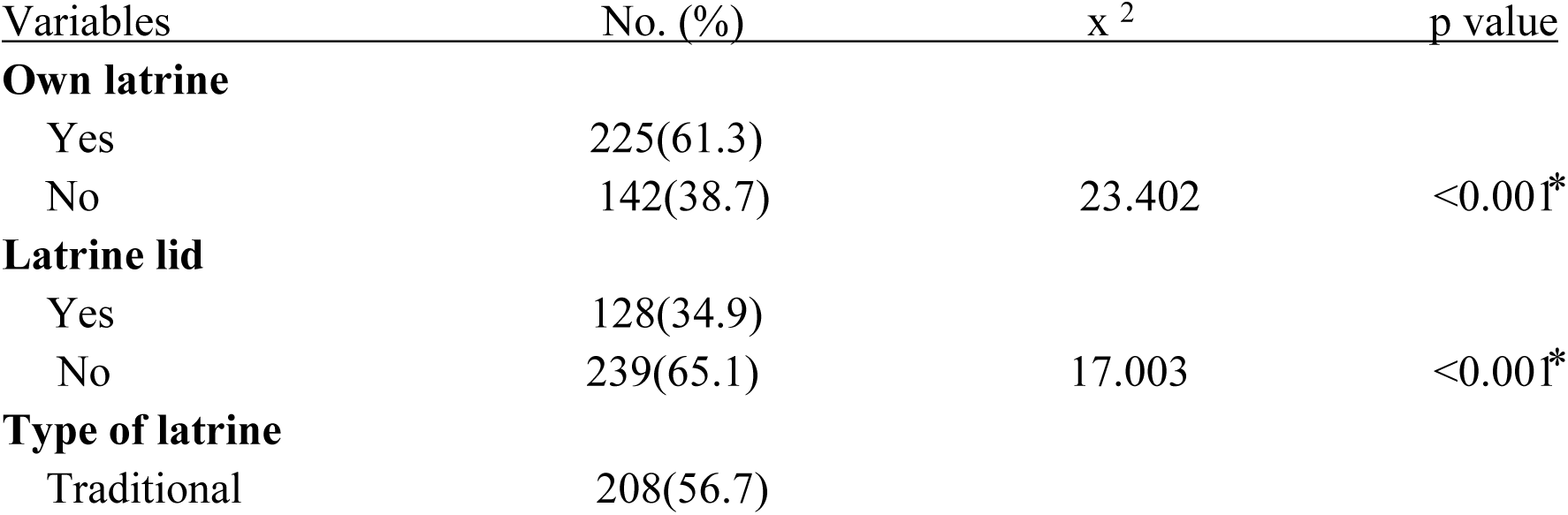
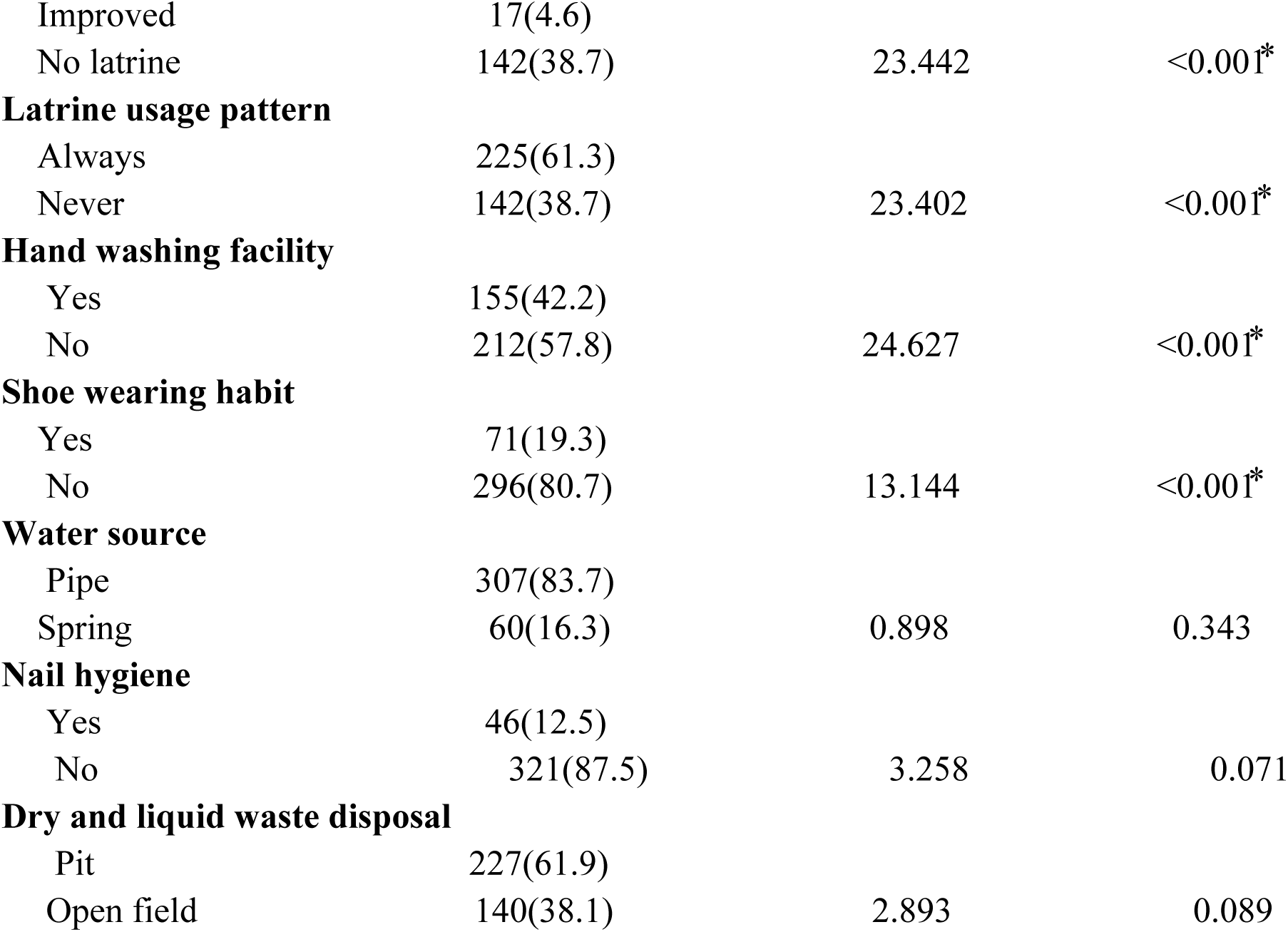
Hygiene and Sanitation-Related Characteristics of the Households, Dawuro zone, Southwestern Ethiopia, 2022/23 (n=367).

### 3.3. Prevalence of Hookworm infection and Anemia

Among the 367 pregnant women who provided stool and blood samples, the prevalence of Hookworm infection and anemia was 10.6% and 31.9% respectively, the mean hemoglobin concentration was 11.33 ± 1.92 g/dl. Three intestinal parasites were identified: trophozoite stages of Giardia lamblia (16.6 %), trophozoite stages of Entamoeba histolytica (5.7 %), and ova of Hookworm (10.6 %) significantly associated with anemia (p < 0.05). Hookworm infection and anemia were most common in women aged 20–24 years (4.4 %) and 25–29 years (2.7 %), although the association with age and trimester was not statistically significant (p > 0.05). Anemia severity was classified as mild (1.1 %), moderate (6.3 %), and severe (3.3 %), with a significant association to hemoglobin level (p < 0.05). Significant associations (p < 0.05) were observed between Hookworm infection and anemia prevalence and several factors, including: educational status, occupational status, residence, low family income and large household size, few ANC visits, third trimester of pregnancy, history of blood loss in last delivery, birth interval, home delivery, lack of iron and folate supplementation, malaria infection in the past year, presence of intestinal parasites, and RBC morphology(normocytic-normochromic)(Table 3).

**Table 3:**
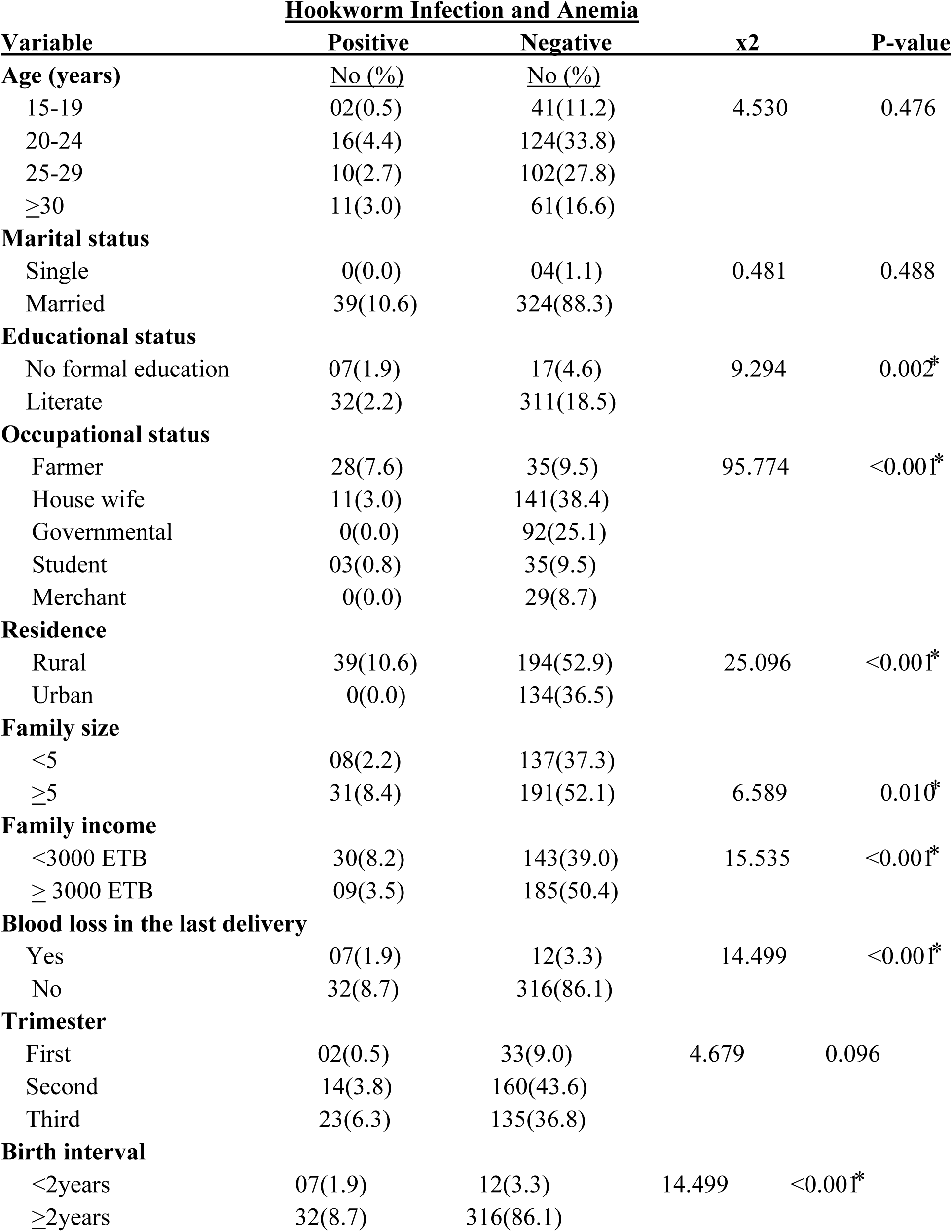
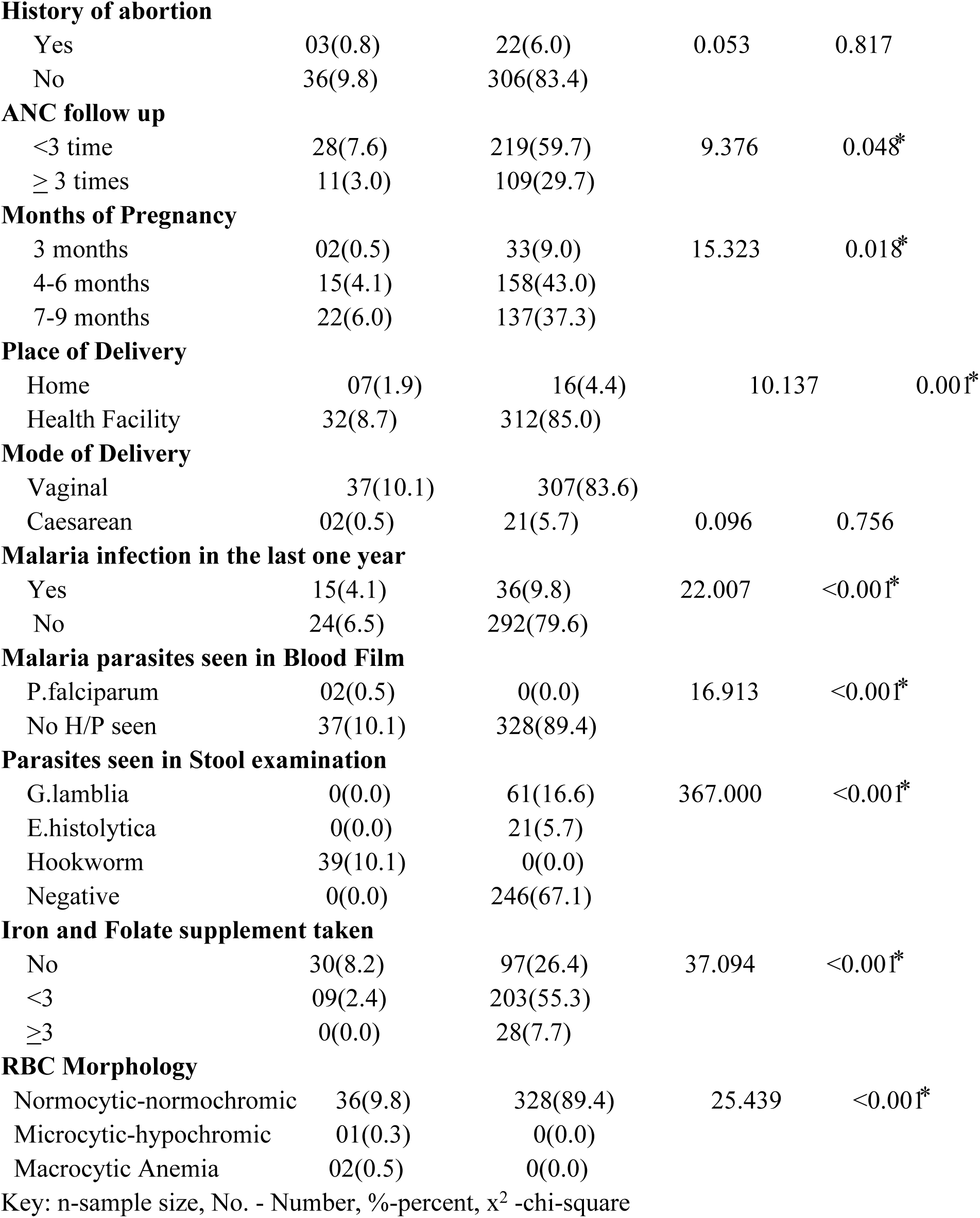
Prevalence of Hookworm infection and Anemia among pregnant women attending ANC in Dawuro zone, Southwest Ethiopia, 2022/23 (n=367)

### 3.4 Severity of Anemia and Associated Factors

The stratification of anemia severity revealed that moderate anemia was the most common (18.5 %), followed by mild (10.6 %) and severe anemia (3.3 %). These findings are consistent with WHO reports indicating that mild to moderate anemia is widespread among pregnant women in low-income countries due to poor nutritional status and parasitic infections like Hookworm. The association between gravidity and anemia severity showed that primi gravida women had significantly higher rates of moderate anemia (6.8 %) compared to multigravida counterparts. This may reflect the physiological and nutritional demands in first pregnancies, compounded by inadequate prenatal supplementation or low iron reserves. Similarly, parity was significantly associated with anemia severity, with para two and para >3 women predominantly showing moderate anemia. This reinforces that repeated pregnancies without adequate spacing or nutrition replenishment increase anemia risk (Table 4).

**Table 4:**
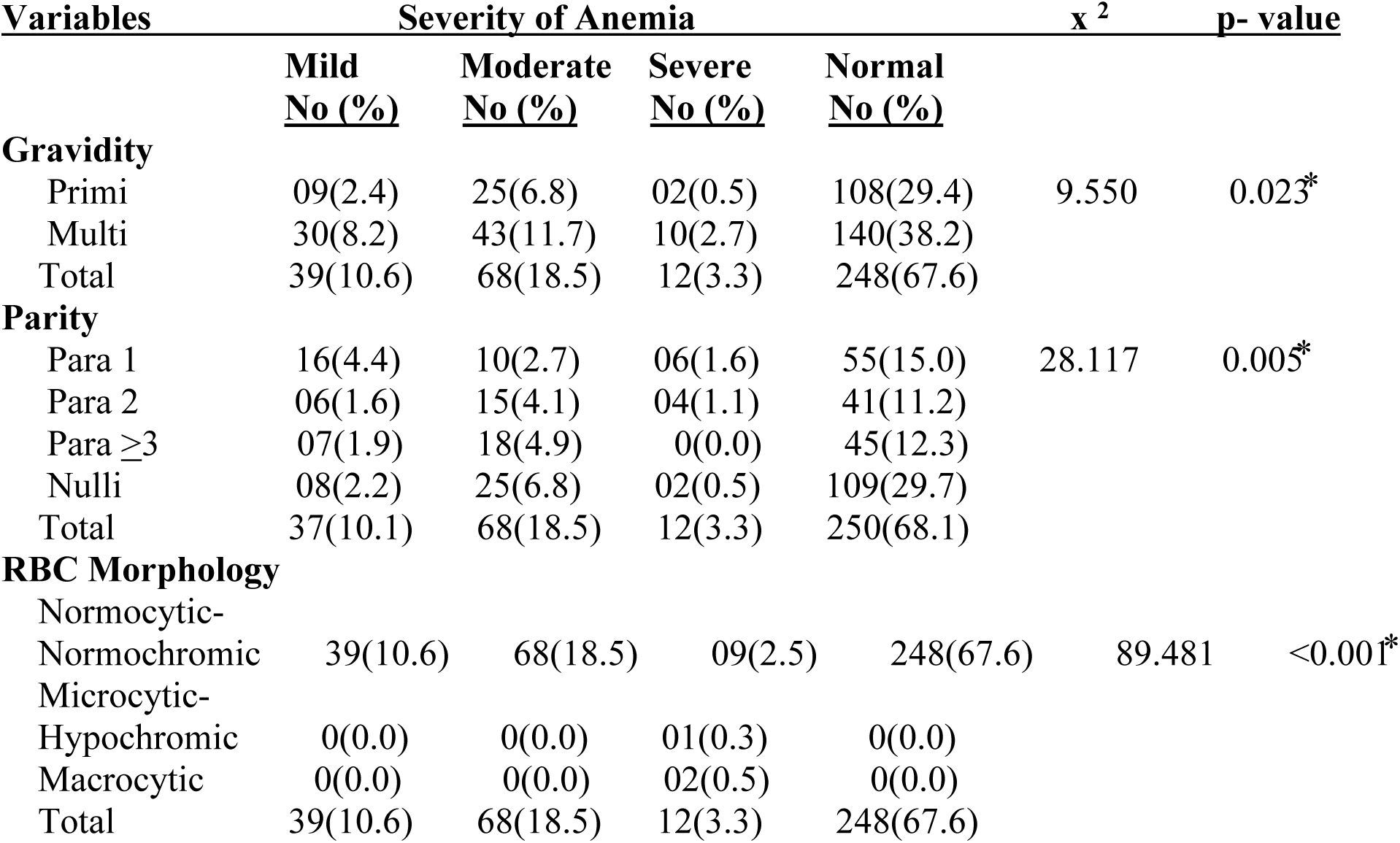
Severity of Anemia among pregnant women attended ANC in Dawuro zone, Southwestern Ethiopia, 2022/23 (n=367)

| Variables | Severity of Anemia |  |  |  | x<sup>2</sup> | p-value |
| --- | --- | --- | --- | --- | --- | --- |
|  | Mild<br>No (%) | Moderate<br>No (%) | Severe<br>No (%) | Normal<br>No (%) |  |  |
| **Gravidity** |  |  |  |  |  |  |
| Primi | 09(2.4) | 25(6.8) | 02(0.5) | 108(29.4) | 9.550 | 0.023* |
| Multi | 30(8.2) | 43(11.7) | 10(2.7) | 140(38.2) |  |  |
| Total | 39(10.6) | 68(18.5) | 12(3.3) | 248(67.6) |  |  |
| **Parity** |  |  |  |  |  |  |
| Para 1 | 16(4.4) | 10(2.7) | 06(1.6) | 55(15.0) | 28.117 | 0.005* |
| Para 2 | 06(1.6) | 15(4.1) | 04(1.1) | 41(11.2) |  |  |
| Para ≥3 | 07(1.9) | 18(4.9) | 0(0.0) | 45(12.3) |  |  |
| Nulli | 08(2.2) | 25(6.8) | 02(0.5) | 109(29.7) |  |  |
| Total | 37(10.1) | 68(18.5) | 12(3.3) | 250(68.1) |  |  |
| **RBC Morphology** |  |  |  |  |  |  |
| Normocytic-Normochromic | 39(10.6) | 68(18.5) | 09(2.5) | 248(67.6) | 89.481 | <0.001* |
| Microcytic-Hypochromic | 0(0.0) | 0(0.0) | 01(0.3) | 0(0.0) |  |  |
| Macrocytic | 0(0.0) | 0(0.0) | 02(0.5) | 0(0.0) |  |  |
| Total | 39(10.6) | 68(18.5) | 12(3.3) | 248(67.6) |  |  |

### 3.5. Red Blood Cell Morphology and Anemia Type

Anemia classification by RBC morphology showed that normocytic-normochromic anemia dominated all severity levels. This pattern is typical of anemia of chronic disease and early iron deficiency. However, among those with severe anemia, cases of microcytic-hypochromic and macrocytic types were also observed, indicating iron deficiency and vitamin B12 or folate deficiency, respectively. These findings underscore the need for routine hematologic profiling in ANC to tailor interventions (Table 4).

### 3.6. Factors Associated with Prevalence of Hookworm infection and Anemia

In the bivariate analysis, multiple factors showed a statistically significant association (p < 0.05) with the prevalence of Hookworm infection and anemia. These included: educational and occupational status, residence, family size and income, latrine-related factors (availability, type, usage, lid presence, and nearby hand washing facilities), personal hygiene (shoe-wearing habit), clinical and obstetric variables (blood loss, birth interval, antenatal care follow-up, gravidity, parity, gestational age, place of delivery, malaria history, malaria species seen in B/F, presence of intestinal parasites, Iron and folate supplementation, and RBC morphology). However, in the multivariate logistic regression, low family income (< 3000 ETB), no latrine access, no hand washing facilities near latrine, not wearing shoes regularly (barefoot habit), fewer ANC visits, advanced gestational age(2nd and 3rd trimesters), gravidity, home delivery, history of blood loss, birth interval, malaria parasite seen in blood film (P.falciparum), and number of Iron and Folate tablets taken were remain significantly associated with prevalence of Hookworm infection and anemia (P<0.05) (Table 5).

**Table 5:**
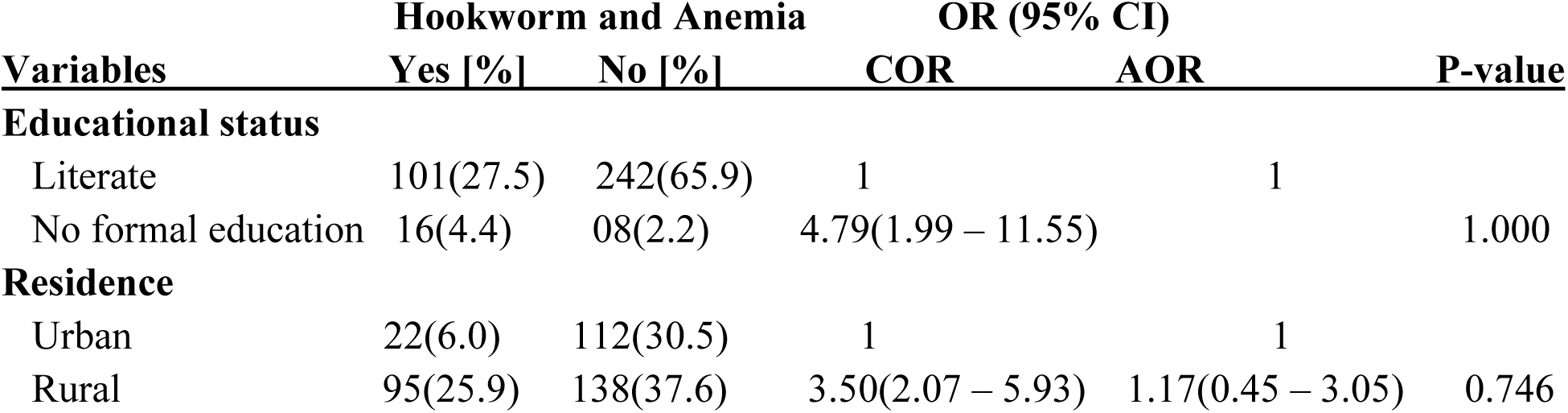
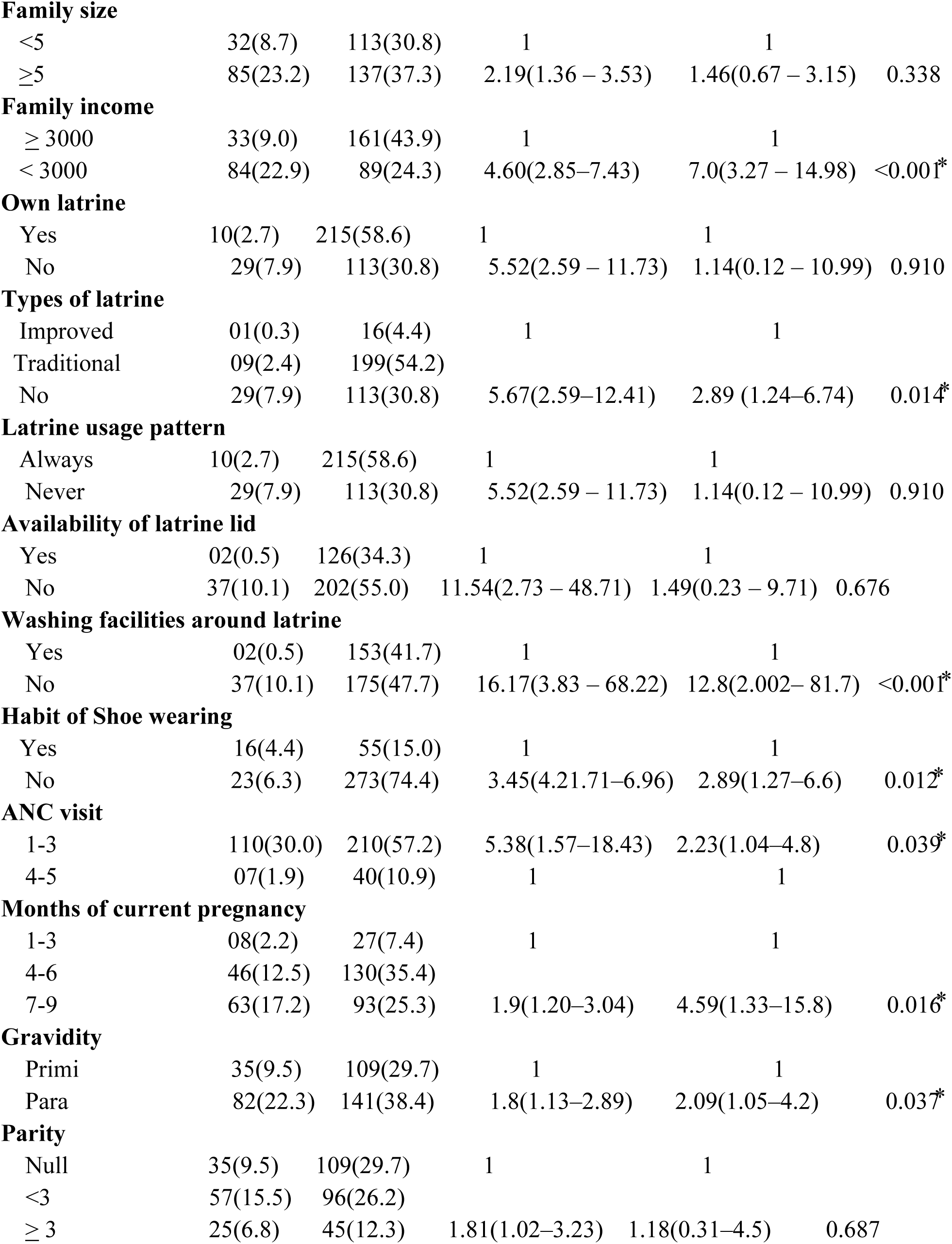
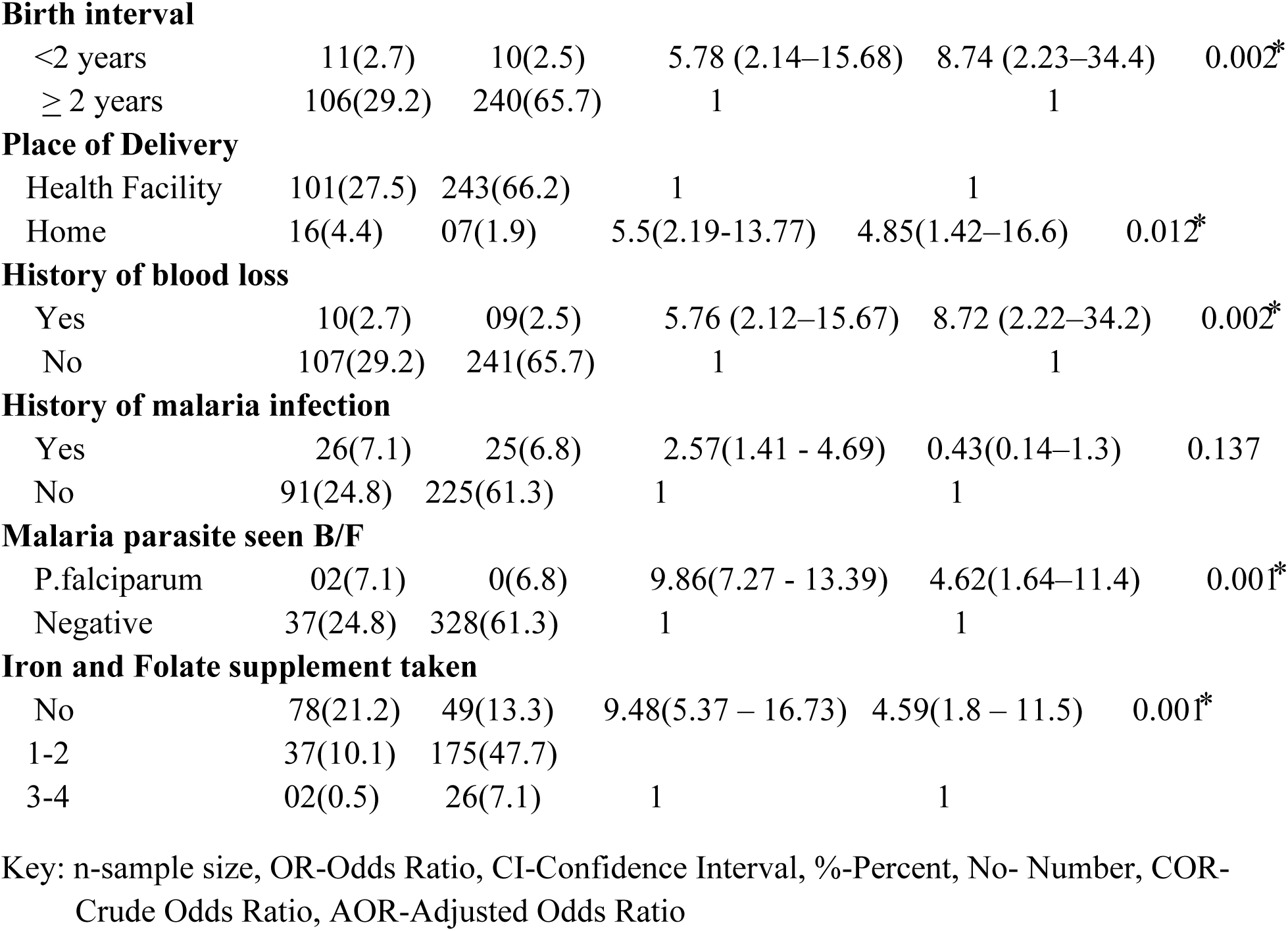
Factors associated with prevalence of Hookworm infection and anemia among pregnant women visited ANC in Dawuro zone, South west Ethiopia, 2022/23 (n=367)

Those pregnant women did not took adequate supplements were 4.6 times more likely to be anemic than those supplemented adequately [AOR=4.59, 95% CI (1.83- 11.53)]. Pregnant mothers in second and third trimester were 5.5 times more likely to be anemic as compared to pregnant mothers in first trimester[AOR=5.534, 95% CI (1.58–19.384)] indicating advanced pregnancy stages increase anemia risk. Hookworm and blood loss [AOR=8.72, 95% CI (2.22- 34.18)] strong evidence that Hookworm-infected mothers experience higher blood loss. Barefoot Habit [AOR=2.89, 95% CI (1.27- 6.61)], confirms walking barefoot significantly increases risk of Hookworm infection and related anemia (Table 5).

## 4. DISCUSSION

### 4.1. PREVALENCE OF HOOKWORM INFECTION AND ANEMIA

The present study assessed the prevalence and associated factors of Hookworm infection and anemia among pregnant women attending antenatal care in Dawuro zone, Southwest Ethiopia. The findings demonstrated a significant burden of Hookworm infection (10.6%), and anemia (31.9 %), highlighting a critical public health challenge in the study area. A total of three intestinal parasites were identified in participants stool sample, namely trophozoite stage of G.lamblia 16.6%, trophozoite stage of E.histolytica 5.7%, and ova of Hookworm 10.6% significantly associated with prevalence of anemia among the study participants (P<0.05). Our study confirms a strong association between Hookworm infection and anemia, consistent with previous findings from East Nepal, where helminthic infection was significantly associated with anemia (46.5 %) among pregnant women [10]. Similarly, research conducted in Kenya revealed that higher Hookworm burden during early pregnancy correlates with increased risk of moderate to severe anemia [11], especially during the first ANC visit, aligning with our finding where low ANC visits significantly increased the odds of Hookworm-anemia co-infection (AOR=2.23,95%CI:(1.04–4.8). This supports the hypothesis that inadequate early antenatal care limits timely diagnosis and treatment of parasitic infections. In Nigeria 35.8% [12], Ghana 45% [13], Uganda 45% [14], Vietnam 78.15% [15], Andhra Pradesh 78.14% [16], Dabat Primary Hospital and Debark General Hospital, Northern Ethiopia 21.4% [17], Dembecha district, Northwest Ethiopia 32% [18], Southwest Ethiopia 59.3% [19], Mecha district, Oromia 14.2% [20],West Gojjam 18.8% [21] had shown higher prevalence of Hookworm infection than the present study. Also the studies conducted among pregnant women in East Nepal 58.9 %( 10), Nairobi Kenya 36.2%(11), south India 51% (35), Southeast Nigeria 40.4% (36), South Africa 57.3% (37), west Algeria 40.08% (38), West Arise zone, Oromia, Ethiopia 38.6% (39) had shown higher prevalence of anemia than the present study. This might be geographic difference, time gap where those studies were conducted, socio demographic and economic conditions of the study participants.

The studies done in some parts of Ethiopia like Northwest Ethiopia 5.5% [20], Hossana 7.0% [22], Addis Ababa 1.3% [23], Felege Hiwot Referral Hospital 1.3%[24],Yirgalem General Hospital 2.7% [25], Gondar Hospital 4.7%[26], and Kenya 3.9%[27] had shown lower prevalence of Hookworm infection among pregnant women than the present study. The high prevalence of Hookworm infection among pregnant women in this study area might be due to the low socioeconomic and educational status of pregnant women, no habit of wearing shoes, and the level of awareness about the transmission, prevention and control of Hookworm infection.

This finding is almost consistent with the study conducted in Uganda 11% (28), Southern Nigeria 11.4% (29), Wondo Genet 11.2% (30), and Kasoa Polyclinic, Ghana 10.3% (31). This might be due to similarity of geographic area, socio demographic, and the level of awareness about the prevention and control of Hookworm infection.

Key socio-demographic factors such as low educational status, rural residence, large family size, and low household income were significantly associated with co-occurrence of Hookworm infection and anemia. These findings are aligned with studies from Eastern Wollega, Oromia, and Western Ethiopia, which reported that illiteracy, low income, and lack of sanitation infrastructure significantly contributed to parasitic infections and anemia (32, 33). Our adjusted analysis confirmed that pregnant women from households earning less than 3000 ETB/month were seven times more likely to have Hookworm infection and anemia (AOR=7.0, 95% CI: 3.27–14.98). The role of education and residence was prominent. Women with no formal education had higher odds of infection, aligning with studies in Nepal and Eastern Wollega (10, 32), which reported higher intestinal parasite prevalence among illiterate mothers. Although our adjusted analysis found education was non-significant, the crude association and supporting literature suggest education still plays a critical role in disease prevention via knowledge and hygiene practices. Environmental sanitation was also significant. Absence of improved latrines, non-use of latrines, and lack of handwashing facilities were strongly linked with co-infection. This is in line with evidence from Eastern Wollega and Northern Tigray (32, 34), where open defecation and absence of latrine covers increased helminth exposure. Notably, in our study, lack of handwashing facilities had a 12.8 times higher likelihood of infection (AOR=12.8, 95% CI: (2.002– 81.7), underlining the critical importance of WASH (water, sanitation, and hygiene) interventions. In terms of maternal behavior, not wearing shoes was associated with higher odds of Hookworm infection (AOR=2.89, 95% CI: (1.27–6.6), reflecting the fecal–soil–skin transmission route of Hookworm. This is supported by numerous studies across Sub-Saharan Africa, including in Nigeria and Ethiopia (36, 40), which link barefoot walking with soil-transmitted helminths. Several obstetric factors also influenced infection. Pregnant women in their third trimester were significantly more likely to suffer from co-infection (AOR=4.59, 95% CI: (1.33–15.8), consistent with findings from Northwest Ethiopia (21), South Africa (37), West Algeria (38), where anemia prevalence peaked in later trimesters. Additionally, multi gravidity, birth intervals <2 years, and home delivery increased the likelihood of infection, corroborating studies from Western Ethiopia and Nigeria (33, 36), which link these factors with nutritional depletion and limited health service access. Our findings reinforce the role of nutritional supplementation, particularly iron and folate, in preventing anemia. Pregnant women who did not take iron and folate had significantly higher odds of anemia (AOR=4.59, 95% CI: (1.8–11.5), a finding echoed in studies from Northwest Ethiopia (21) and Northern Tigray (34). However, similar to results from Gondar University Hospital (26) and India (35), supplementation alone was insufficient in preventing anemia in some cases, highlighting the need for holistic nutritional, parasitic, and behavioral interventions. Interestingly, a history of malaria and blood loss was both associated with increased anemia risk. Although malaria infection did not remain significant after adjustment (p>0.05), the presence of P. falciparum parasites did (AOR=4.62, 95% CI: (1.64–11.4), suggesting that co-infection exacerbates anemia severity a trend also seen in Sub-Saharan Africa including Ethiopia (40).

## Conclusion

This study demonstrated that Hookworm infection and anemia remain significant public health problems among pregnant women in Dawuro zone, Southwest Ethiopia. Co-infection was found to be influenced by a multitude of factors, including low ANC attendance, poor hygiene and sanitation practices, parasitic co-infection, late pregnancy trimester, lack of footwear, and absence of iron supplementation. The findings are in agreement with a range of studies conducted across Ethiopia and other developing countries, reaffirming the need for integrated maternal health interventions. To mitigate the dual burden of Hookworm and anemia during pregnancy, early ANC visits, improved sanitation, health education, and nutritional support must be prioritized. A coordinated, multisectoral approach is critical to achieving improved maternal and fetal health outcomes.

## Data Availability

The data used to support the findings of this study are available from the corresponding author on reasonable request.

## Author Contributions

Conceptualization: Getachew Teshome, Berhanu Effa, Asrat Chernet.

Data curation: Getachew Teshome.

Formal analysis: Getachew Teshome.

Investigation: Asrat Chernet, Getachew Teshome.

Methodology: Getachew Teshome, Temesgen Weldemedhin, Mengistu Geribo

Project administration: Getachew Teshome.

Resources: Getachew Teshome.

Software: Getachew Teshome, Berhanu Effa.

Supervision: Getachew Teshome, Berhanu Effa, Temesgen Weldemedhin.

Validation: Getachew Teshome, Asrat Chernet, Mengistu Geribo.

Visualization: Getachew Teshome, Berhanu Effa, Temesgen Weldemedhin.

Writing – original draft: Berhanu Effa, Getachew Teshome.

Writing – review & editing: Getachew Teshome, Berhanu Effa, Asrat Chernet, Temesgen Weldemedhin, Mengistu Geribo.

## Acknowledgments

We thank Wolaita Sodo University for its support during the research activities and funding this research. We also extend our thanks to the Dawuro zone health department, study participants, data collectors, supervisors, and colleagues for their cooperation and support during the research activities.

